# US women screen at lower rates for both cervical and colorectal cancers than a single cancer

**DOI:** 10.1101/2021.11.28.21266963

**Authors:** Diane M Harper, Melissa Plegue, Masahito Jimbo, Ananda Sen, Sherri Sheinfeld Gorin

**Affiliations:** Department of Family Medicine, University of Michigan, Ann Arbor, MI, USA; Department of Obstetrics and Gynecology, University of Michigan, Ann Arbor, MI, USA; Department of Women’s and Gender Studies, University of Michigan, Ann Arbor, MI, USA; Department of Biostatistics, University of Michigan, Ann Arbor, MI, USA; Department of Family and Community Medicine, University of Illinois at Chicago, Chicago, IL, USA

**Keywords:** colorectal cancer screening, cervical cancer screening, self-screening, home-based screening, women, adult, dual screening

## Abstract

**Introduction:** Using screen counts, women 50-64 yo have lower cancer screening rates for cervical and colorectal cancers compared to all other age ranges. The primary aim of this paper is to present cervical cancer and CRC screenings per woman and determine the predictors of being up-to-date for both.

**Methods:** We used the Behavioral Risk Factor Surveillance System (BRFSS), an annual survey to guide health policy in the US, to explore the up-to-date status of dual cervical and colorectal cancer screening for women 50-64 yo. We categorized women into four mutually exclusive categories: up-to-date for dual screening, each single-screen, or neither screen. Multinomial multivariate regression modeling was used to evaluate the predictors of each category.

**Results:** Among women ages 50-64 yo, dual screening was reported for 58.7% (57.6-59.9), cervical cancer screening alone (27.0% (25.9-28.1), CRC screening alone (5.3% (4.8-5.8), and neither screen (9% (8.4-9.6). Age, race, education, income, and chronic health conditions were significantly associated with dual screening compared to neither screen. Hispanic women compared to non-Hispanic White women were more likely to be up-to-date with cervical cancer screening than dual screening (aOR 1.38 (1.08, 1.77). By comparison to younger women, those 60-64 years are significantly more likely to be up-to-date with CRC screening than dual screening (aOR 1.75 (1.30, 2.34)).

**Conclusions:** Screening received by each woman shows a much lower rate of dual screening than prior single cancer screening rates. Addressing dual screening strategies rather than single cancer screening programs for women 50-64 years may increase both cancer screening rates.

**Graphical Abstract:** Screening rates differ by calculation approach. A better population metric for cancer prevention is to consider the screens each woman has received. Our data show much lower cervical cancer and CRC screening rates than a single screen calculation.

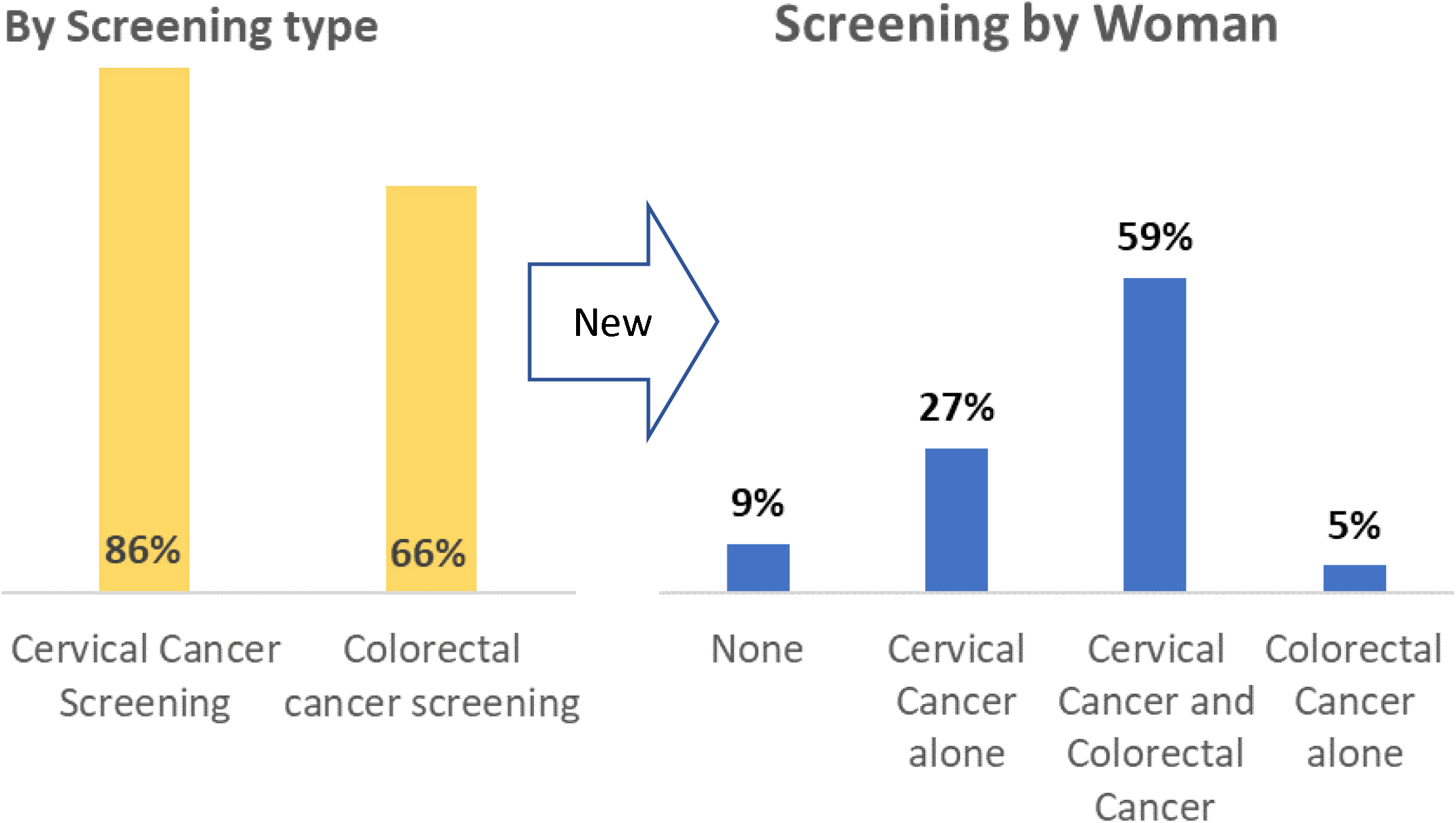

**Tweet:** Women 50-64 years old lack dual cervical and colorectal cancer screening. Women who screen for only one cancer of the two, screen for cervical cancer almost five times more often than colorectal cancer.

**Lay Summary:** Routine cervical and colorectal cancer (CRC) screenings can detect early, treatable, curable cancers. Using the responses to the 2018 Behavioral Risk Factor Surveillance Survey (BRFSS) we approached prevention by what the woman has received rather than by how many screens were done. We showed that women 50-64 years old were only up-to-date for both cervical and CRC screening 59% of the time, whereas, by traditional screening calculations, our data showed 86% of women were up-to-date for cervical cancer screening and 64% were up to date with CRC screening, irrespective of any other cancer screenings.

## Introduction

One of the primary goals of Healthy People 2030 is to improve cancer screening behaviors with evidence-based screening and prevention strategies (**HP2030-CR02 (1)**). On the population level, the Behavioral Risk Factor Surveillance System (BRFSS) survey collects data on cancer screenings by cancer type, by screening method and by the time from their last screening (**BRFSS codebook 2019 (2)**). BRFSS is a common database used to report single cancer screening rates, such as cervical cancer or colorectal cancer. Never has the perspective of screening by woman been published reporting the patterns of screening for both cervical cancer and colorectal cancer (CRC) (*dual screening*) among women 50-65 years old.

Colorectal cancer (CRC) screening has never approached the Healthy People goals, but up-to-date screening has shown a steady increase, regardless of screening method (**NCI Progress Report 2020 CRC (3)**). Most recently, however, BRFSS showed a significant gap in CRC screening among women 50-64 years of age with self-reported rates as low as 50% for the youngest age-appropriate women (**Joseph 2020 (4)**). Unlike CRC screening, cervical cancer screening prevalence peaked at the turn of the 21^st^ century and has since been declining year over year (**NCI Progress Report 2020, summary table (5)**). Like CRC screening, the lowest rates of cervical cancer screening are among women who were 50-64 years old (**Harper 2020 (6)**).

Both cervical and colorectal cancer generally do not have symptoms until a very late stage where cure is not possible (**WHO 2017 (7)**), making early screening, even self-screening **(El Khoury 2021 (8), Bakr 2020 (9), Jaklevic 2020 (10)**), an important health imperative.

The primary aim of this paper is to present cervical cancer and CRC screenings per woman 50-64 years old and determine the predictors of being up-to-date in <u>both</u> cervical cancer and CRC screening (*dual-screened*) compared to each single screen.

## Methods

### Survey Data

BRFSS is an annual, state-based, cross-sectional telephone survey that US state health departments conduct monthly using landline telephones and cellular telephones using a standardized questionnaire developed by the Centers for Disease Control and Prevention (CDC). Individual residents self-report their health risk behaviors as well as their completion of clinical preventive services. CRC screening is defined as using a home kit for blood stool tests including fecal occult blood test (FOBT) or fecal immunochemical test (FIT) or office-based procedures including sigmoidoscopy or colonoscopy. The interval options since the last screening include 1) within the past year (anytime less than 12 months ago), 2) within the past 2 years (1 year but less than 2 years ago), 3) within the past 3 years (2 years but less than 3 years ago), 4) within the past 5 years (3 years but less than 5 years ago), and 5) five or more years ago. The responses for cervical cancer screening are based on office-based testing for HPV, Pap test, or both. The interval options since the last screening were identical to those for CRC screening. The BRFSS dataset available for our analysis was 2018.

### Screening outcomes

Cervical cancer screening was considered up-to-date if the respondent reported a Pap within the past 3 years, an HPV test within the past 5 years, or a Pap and HPV test within the past 5 years (**USPSTF 2018 (11)**). We included the five-year screening interval in our outcome measure of cervical cancer screening. For colorectal cancer screening, up-to-date screening included FOBT or FIT within the past year, a sigmoidoscopy within the last 5 years, or a colonoscopy within the past 10 years, in accordance with the USPSTF guidelines (**USPSTF 2016 (12), USPSTF 2021 (13)**). To examine up-to-date screening for both cancers, we grouped women in four mutually exclusive screening categories: those who were up-to-date for both screens (dual screening), those up-to-date with a single screen (cervical cancer screen only or CRC screen only), and those who denied completing either screen.

### Survey respondents

We included women aged 50-64 years with no reported history of uterine or cervical cancer and who had not had a hysterectomy. We also excluded those with reported colon or rectal cancers.

### Statistical analysis

Data were summarized using proportions, unweighted and weighted, with 95% confidence intervals. The dataset was analyzed using weighted sampling methods to ensure valid inferences from the responding sample to the general population, correcting for non-response and non-coverage biases (**StataCorp, 2017 (14), BRFSS Sample Weights 2018 (15))**. Composite up-to-date screening prevalences were summarized across all screening methods for each cancer screen. For instance, a woman was up-to-date with CRC screening if she had reported a FIT or FOBT test within the past 1 OR a sigmoidoscopy within the past 5 years OR a colonoscopy within the past 10 years. Likewise, a woman was up-to-date with cervical cancer screening if she had reported a Pap test within the past 3 years OR a combined Pap and HPV test within the past 5 years OR an HPV test within the past 5 years. Women with complete screening status were included in the analysis.

Multinomial logistic regression was used to evaluate the associations with the predictors of screening outcomes. Predictors of interest were age, race, education, income, marital status, occupational status, geolocation, and the presence of any chronic health condition as found to be pertinent in past work (**Harper 2020 (6)**). All tests of statistical significance used a p-value threshold of 0.05. This retrospective study was exempt from the IRB.

## Results

There were 67,473 women identified between the ages of 50-64 years from which 40,511 met the inclusion and exclusion criteria and reported complete screening data for CRC and cervical cancers in the BRFSS 2018 survey (**Supplemental figure 1 CONSORT diagram**). Weighted prevalences for the sociodemographic data are presented in **Table 1**, where the largest proportion of women were urban, married, attended at least some college, of White race, and employed.

**Table 1.**
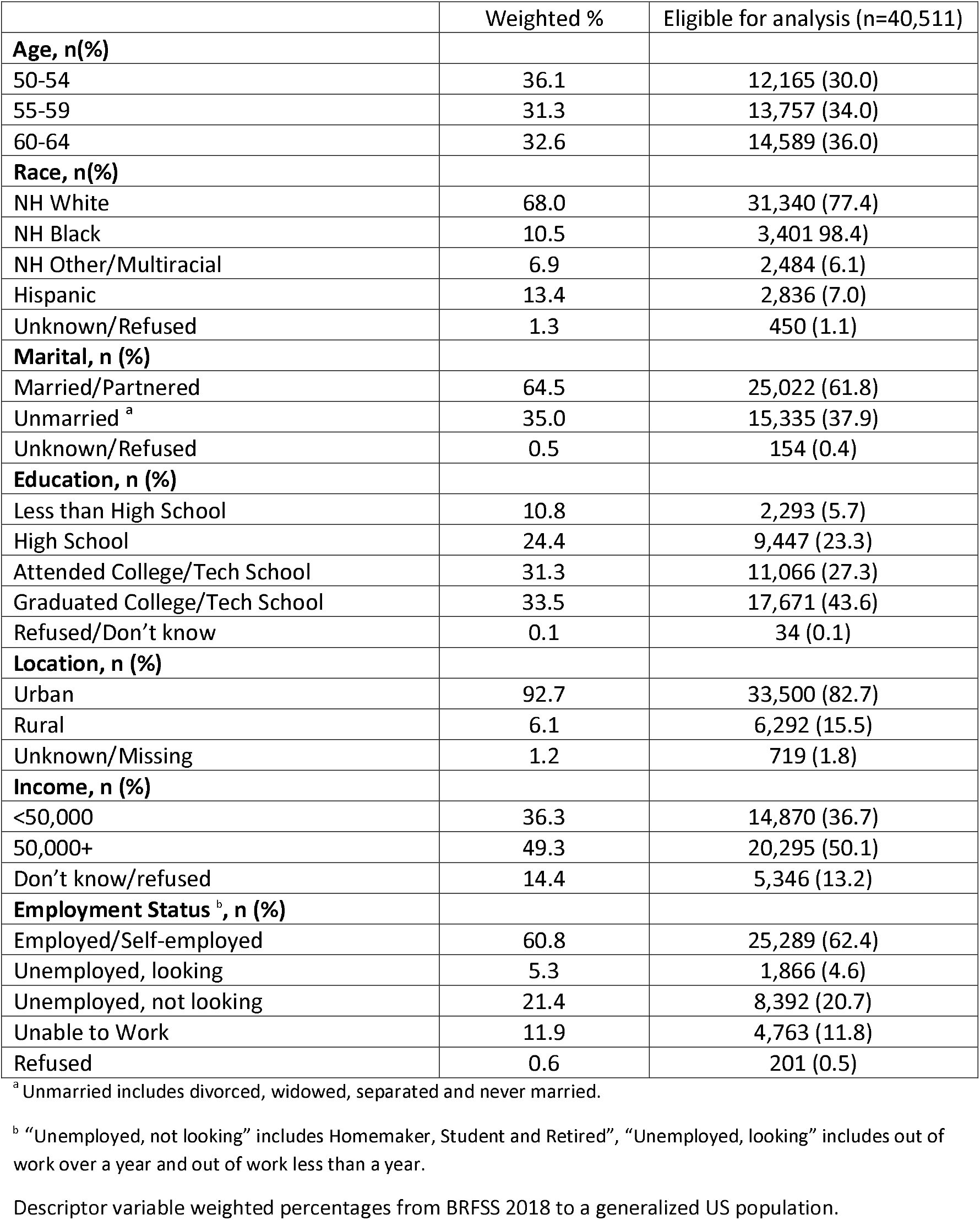
Demographic characteristics of the study population

**Table 2** presents mutually exclusive up-to-date screening categories by screening modality. The weighted prevalence for women with dual CRC and cervical cancer screening was **58.7%** (57.6%, 59.9%), n=24,678), for women with neither screen was **9.0%** (8.4%, 9.6%), n=3,560), for women with only cervical cancer screening was **27.0%** (25.9%, 28.1%), n=10,051), and for women with only CRC screening was **5.3%** (4.8%, 5.8%)], n=2,222). The tests used by the majority of women for CRC and cervical cancer screening, respectively, were colonoscopy and a Pap test (**54.6**% (53.2%, 55.9%)). Women who chose only one screen chose cervical cancer screening 5 times more often than colorectal cancer screening (27% vs. 5.3%). By traditional screening calculations, cervical cancer screening was 86% (85.4-86.1) and CRC screening was 66% (65.9-66.8) (see **Graphic Abstract**).

**Table 2.**
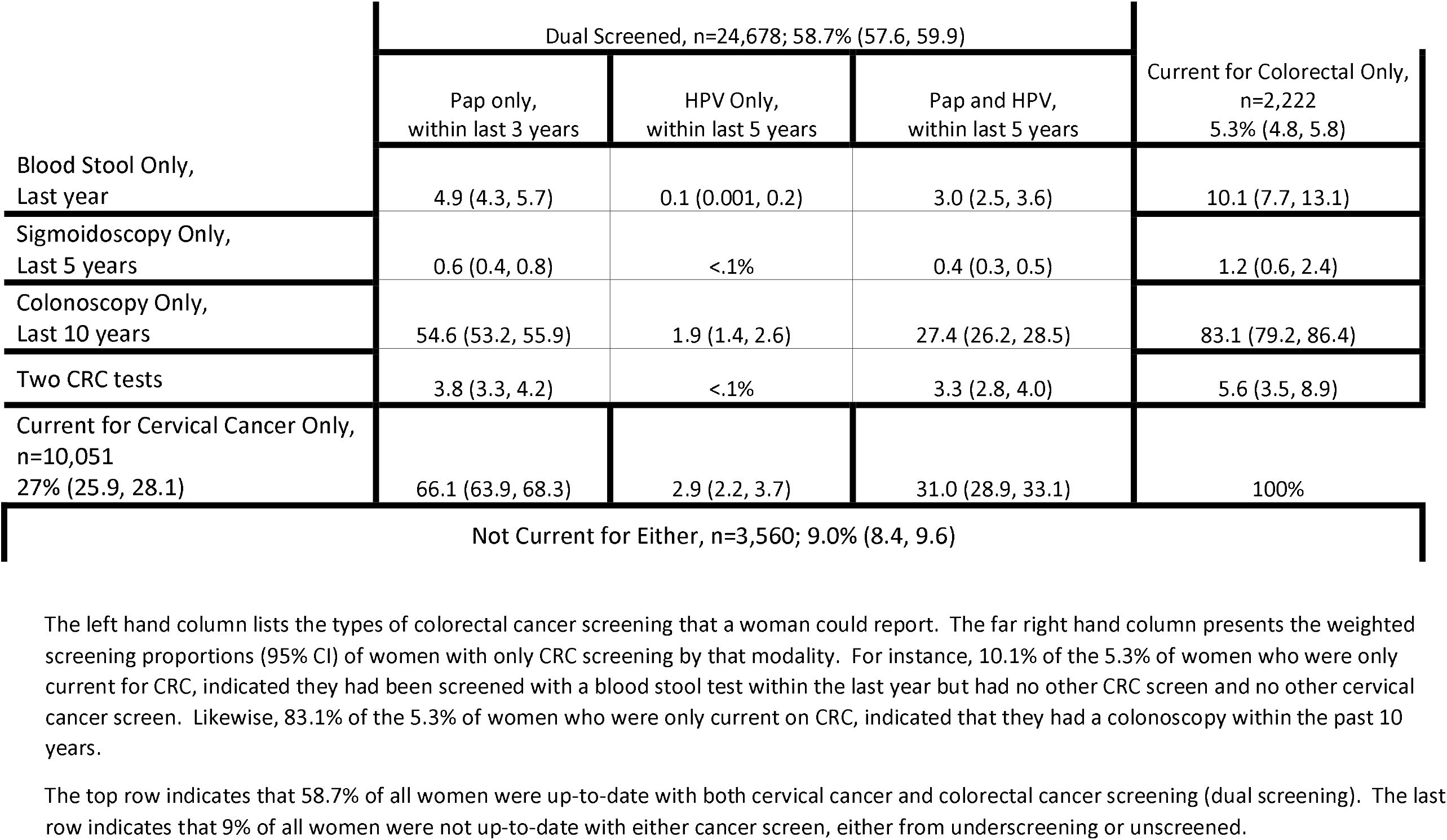
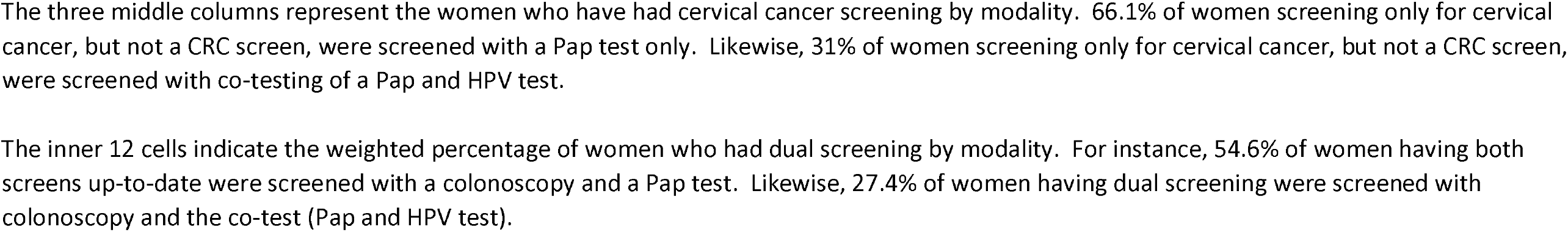
Up to date screening status by screening modality

|  | Dual Screened, n=24,678; 58.7% (57.6, 59.9) |  |  | Current for Colorectal Only,<br>n=2,222<br>5.3% (4.8, 5.8) |
| --- | --- | --- | --- | --- |
|  | Pap only,<br>within last 3 years | HPV Only,<br>within last 5 years | Pap and HPV,<br>within last 5 years |  |
| Blood Stool Only,<br>Last year | 4.9 (4.3, 5.7) | 0.1 (0.001, 0.2) | 3.0 (2.5, 3.6) | 10.1 (7.7, 13.1) |
| Sigmoidoscopy Only,<br>Last 5 years | 0.6 (0.4, 0.8) | <.1% | 0.4 (0.3, 0.5) | 1.2 (0.6, 2.4) |
| Colonoscopy Only,<br>Last 10 years | 54.6 (53.2, 55.9) | 1.9 (1.4, 2.6) | 27.4 (26.2, 28.5) | 83.1 (79.2, 86.4) |
| Two CRC tests | 3.8 (3.3, 4.2) | <.1% | 3.3 (2.8, 4.0) | 5.6 (3.5, 8.9) |
| Current for Cervical Cancer Only,<br>n=10,051<br>27% (25.9, 28.1) | 66.1 (63.9, 68.3) | 2.9 (2.2, 3.7) | 31.0 (28.9, 33.1) | 100% |
| Not Current for Either, n=3,560; 9.0% (8.4, 9.6) |  |  |  |  |

The left hand column lists the types of colorectal cancer screening that a woman could report. The far right hand column presents the weighted screening proportions (95% CI) of women with only CRC screening by that modality. For instance, 10.1% of the 5.3% of women who were only current for CRC, indicated they had been screened with a blood stool test within the last year but had no other CRC screen and no other cervical cancer screen. Likewise, 83.1% of the 5.3% of women who were only current on CRC, indicated that they had a colonoscopy within the past 10 years.
The top row indicates that 58.7% of all women were up-to-date with both cervical cancer and colorectal cancer screening (dual screening). The last row indicates that 9% of all women were not up-to-date with either cancer screen, either from underscreening or unscreened.
The three middle columns represent the women who have had cervical cancer screening by modality. 66.1% of women screening only for cervical cancer, but not a CRC screen, were screened with a Pap test only. Likewise, 31% of women screening only for cervical cancer, but not a CRC screen, were screened with co-testing of a Pap and HPV test.
The inner 12 cells indicate the weighted percentage of women who had dual screening by modality. For instance, 54.6% of women having both screens up-to-date were screened with a colonoscopy and a Pap test. Likewise, 27.4% of women having dual screening were screened with colonoscopy and the co-test (Pap and HPV test).

The **descriptive analyses** (**Table 3**) showed differences among women across the four mutually exclusive groups. **Age** by five-year intervals showed that the youngest group was least screened for both CRC and cervical cancer (dual screening) (47.9%) as well as CRC screening alone (3.0%). However, this age group had the highest screening for cervical cancer alone (40.0%). Among **races**, Hispanic women had the lowest dual screening (47.0%), the lowest screening for CRC alone (5.2%), and yet had the highest screening for cervical cancer alone (39.2%).

**Table 3.**
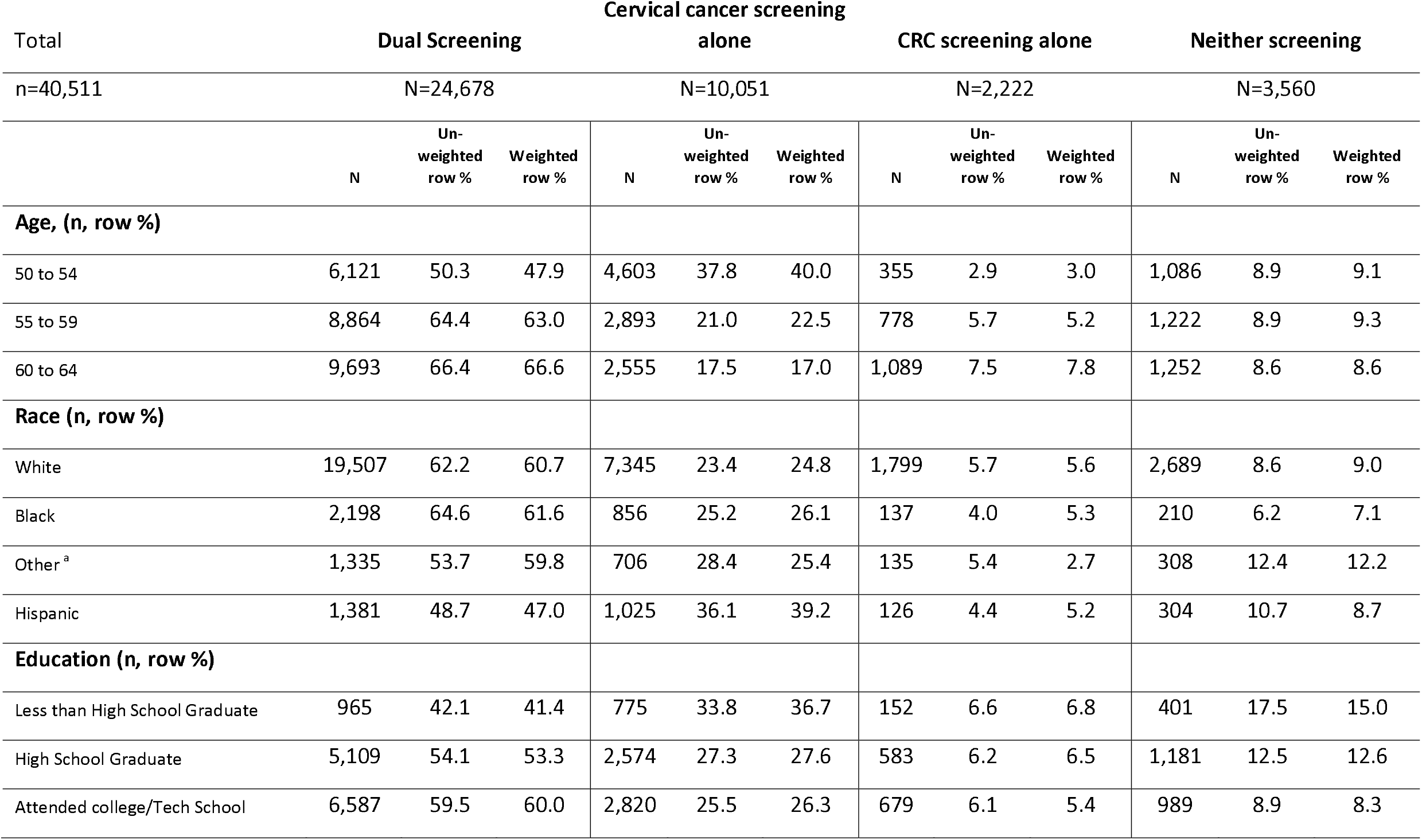
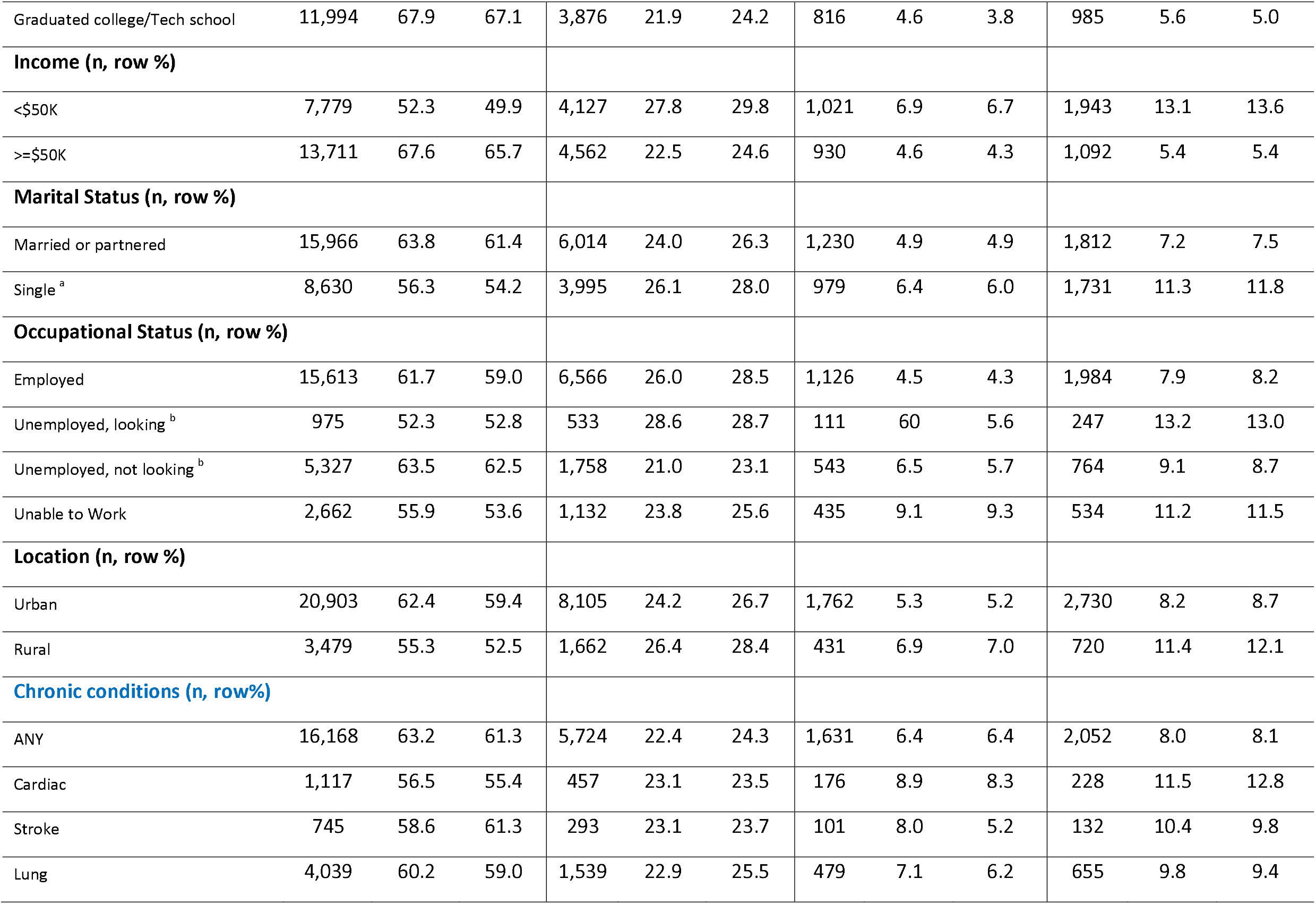
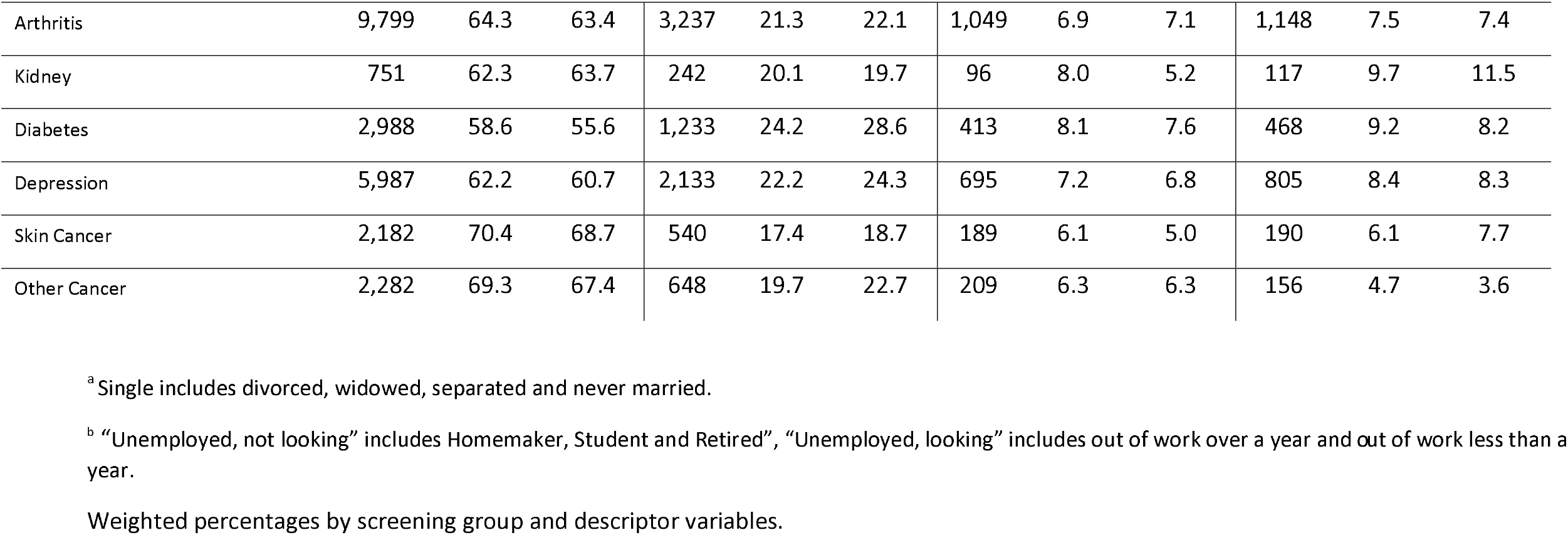
Sociodemographic descriptors by screening category.

Among women of all **education** levels, those with the least education had the lowest dual screening (41.4%), but the most for a single cancer (cervical (36.7%); CRC alone (6.8%)). In addition, the least educated had the highest percentage of not participating in either screen (15%). Conversely, among all **income** levels, those with the highest income reported the greatest dual screening (65.7%) and the lowest single cancer screening (cervical: 24.6%; CRC: 4.3%).

Among **marital** categories, never-married women reported the lowest dual screening (54.2%); and, the highest for the single screens (cervical: 28%; CRC 6%). Unemployed women (homemaker, student, or retired) among all **employment** categories reported the highest dual screening (62.5%), while those living in rural areas reported the lowest dual screening (52.5%). 61% of women with chronic conditions completed dual screening.

For each of the candidate predictor variables, unadjusted multinomial logistic regression results are presented in the supplementary table (**Supplementary Table 1**).

### Multivariate multinomial logistic regression

Multivariate multinomial logistic regression was used to evaluate screening outcomes adjusted for the significant model matched variables of age, race, education, income, and some chronic conditions. Wald tests were used to test the inclusion of the variable in the overall model where marital status (p =0.202), current employment (p =0.189), geographic location (p =0.08), cardiac comorbidities (p =0.12), kidney comorbidities (p =0.11) and lung comorbidities (p =0.64) were no longer significant and, hence, were dropped from the model (**Table 4**).

#### Screening compared to neither screening

Women who graduated from college compared to less than high school graduation, who had the highest income level or had experienced prior cancer had the highest adjusted odds of **completing dual screening** compared to no screening. In addition, women who were older, Black or Hispanic compared to White, and past history of arthritis, diabetes, or depression were also significantly more likely to be up-to-date for **both screens** compared to neither screen.

Hispanic women compared to White had the highest adjusted odds of completing **just the cervical cancer screening** compared to neither screening. In addition, women in the highest income level, who graduated from college compared to less than high school graduation or had a prior cancer were also more than twice as likely to have only cervical cancer screening compared to having no screening. Finally, younger women, Black compared to White, and those with a past history of arthritis, or diabetes were more likely to screen for **cervical cancer alone** than neither screening.

Women with prior cancer had the highest adjusted odds of completing **just the CRC screen** compared to having neither screening. The oldest women, those with the highest income level or those with past history of arthritis were more than two times more likely to have CRC screening alone compared to neither screening. Finally, women with past history of diabetes or depression were more likely to screen for **CRC only** compared to neither screening. Conversely, women who were American Indian, Native Hawaiian, Asian, or multiracial were significantly less likely to be up-to-date for CRC screening compared to neither screening.

#### No or single screen compared to dual screening

Only Hispanic women compared to White women had significantly higher adjusted odds of having **cervical cancer screening alone** compared to dual screening. Alternatively, older women, Black women compared to White, those attending at least some college compared to less than high school, those with higher incomes, or those with arthritis, depression, skin cancer, or other cancers were significantly less likely to have **cervical cancer screening alone** compared to completing both screens.

Only women 60-64 years old compared to younger women had significantly higher adjusted odds for having **CRC only screening** compared to both screens. Women who were American Indian, Native Hawaiian, Asian, or Multiracial, graduated from college, had a higher income, or had past history of skin cancer were significantly less likely to be up-to-date for **CRC screening alone** compared to both screens.

#### Only CRC screening compared to only cervical cancer screening

Older women compared to younger women had the highest adjusted odds of completing **only the CRC screening** compared to cervical cancer screening alone. In addition, women with past history of arthritis or depression had significantly higher adjusted odds of having **only CRC screening** compared to only cervical cancer screening. Women who were American Indian, Native Hawaiian, Asian, or multiracial were less likely to have **only CRC screening** compared to only cervical cancer screening.

## Discussion

### Screening rates much lower in the by-woman analysis

BRFSS’ large scale nationally weighted survey data shows a new finding that the rate of women who screened for both cervix and colorectal cancer is far below the recommended rates for each cancer individually (HP2030 goals cervix: 84.3% and CRC:74.4%) (**HP2030 C09 (16), HP2030-C07 (17)**). Another new finding is that women 50-64 years old screen for cervical cancer alone five times more often than CRC alone. We show that the 50-64 year age range is dynamic with changing cancer screening behaviors from mostly cervical screening at 50 years to mostly CRC screening at 65 years. These results are similar to the population survey study completed in southeast Michigan (**Harper 2021 (18)**).

### Screening Decision Making at Multiple Levels

Age becomes the most interesting factor predicting the divergence of the two cancer screening rates. One hypothesis is that it may be due to the specialty of the physician to whom the woman visits, a gynecologist vs. a PCP (family physician vs. general internist vs geriatrician). A study of mammography orders noted that gynecologists provided care for 15% of the population who had mammograms ordered for ages 50-75 years, a small percentage of the age-appropriate screening population (**Taplin 1994 (19)**). In addition, there is a 90% drop in visits to a gynecologist for prevention-related visits for those who are 45 years or older compared to the 18-44 years old group, with a concomitant increase in PCP visits during this age transition (**Scholle2002 (20)**). This indicates that the **decision level** for the type of cancer screening could be at the *physician le*vel where specialty may influence the cancer screening (cervical vs. colorectal vs. both) offered to the woman.

Another **decision level** for screening could be at the *level of the woman*. The effects on screening by the age divergence could be due to physiologic aging with menopause at an average of 52 years in the US (**OWH (21), Green 2009 (22)**). She may choose not to have any further cervical cancer screenings by vaginal speculum exam due to a variety of possible reasons: painful exams, sexually inactive, no symptoms, or other personal reasons including abuse (**Cadman 2012 (23), Saunders 2021 (24), Gunes 2017 (25)**). She may choose to wait until 65 years to start CRC screening because of health insurance coverage, out-of-pocket costs, and high deductibles, or because of the time off from work necessary to complete the preparation and exam if colonoscopy is the chosen modality for screening.

Decision making about screening could also reside at *the health system* as was evident during COVID when mailed FIT tests were sent to enrollees in response to the drop in cancer screenings (**BCBS 2020 (26), Gupta 2020 (27), Gorin 2020 (28) Van Hoy 2020 (29)**). It is well-established that CRC screening, as recommended by the USPSTF (**USPSTF 2016 (12), USPSTF 2021 (13)**) can be completed by any of six available tests, three of which are home-based: fecal occult blood test (FOBT), fecal immunochemical test (FIT), and multitarget stool DNA (FIT-DNA, Cologuard®). Most recently, the American Cancer Society (ACS) and the USPSTF have recommended primary HPV testing for women 25/30-65 years for cervical cancer screening (**ACS (30), USPSTF 2018 (11)**) as the test that provides the most benefit for the least harms which can also be an at-home test, a well-accepted option (**Maver 2020 (31), Kim 2017 (32), El Khoury 2021 (8**)). Nevertheless, our data show that of those women who only choose one cancer to screen for, cervical cancer will be almost 5 times more likely to be the screen chosen. We hypothesize from our study that If a home-based cervical cancer screening was FDA approved, it could enhance uptake of the already FDA approved CRC home-based testing for women in this vulnerable age range (**Bakr 2020 (9), Jaklevic 2020 (10)**).

Another hypothesis-generating result of this study is that the high rates of a single cancer screening among the least educated may represent screening divorced from their routine primary care. Perhaps the single screen was completed at a local health fair, worksite, or a targeted program for that single cancer (**NBCCEDP 2020 (33), CRCCP 2020 (34**)), achieving the maximum screening rates in this manner, but still below the HP2030 goals. We have shown that women who screen for one cancer do not necessarily screen for other cancers. In prior work, we have shown that having a strong relationship with a primary care physician (PCP) leads to greater participation in dual cervical and colorectal cancer screens (**Harper 2021 (35)**) and we encourage leveraging the power of the PCP-woman dyad to address her multiple competing health needs. We showed that the single screen predictors of self-reported location (urban vs rural) and employment status (**Harper 2020 (6)**) were **not associated with dual screening** in the final multivariate multinominal model. Again, this could support the hypothesis that the relationship between the PCP and woman rather than their location or her insurance type may increase cancer screening rates beyond the single cancer screen.

### Limitations

BRFSS is a cross-sectional self-report survey whose responses are not equivalent to validated medical records or claims data (**St Clair 2017 (36)**). Others have shown that the reported cancer screens could be mistakenly over-estimated by about half of those who did not have a screen, whereas they accurately reflect those who have screened (**Anderson 2019 (37), Bonafede 2019 (38)**). In addition, for cervical cancer screening, likely, women were not aware of the screening modality being used, and hence overestimated Pap alone screening when co-testing may have been done (**Watson 2018 (39))**. For women 50-65 years old, though, co-testing is less common than Pap testing alone mitigating that possible over-estimation (**Watson 2018 (39))**.

Our age range, 50-64 years, was aligned with the screening guidelines at the time of BRFSS 2018 survey. The most recent CRC recommendations by USPSTF move the age of initiation of CRC screening to 45 years, providing a greater potential age gap to be addressed for dual screening. Finally, our work applies to the United States and the health care structure it provides. These results may not apply to other national health care systems.

## Conclusions

We showed that the risk factors associated with dual cancer screening are different from those associated with single cancer screens and that the risk factors for cervical cancer screening only are different from CRC screening only. We showed that women who have dual screening compared to no screening are more likely to have had prior cancer.

## Supporting information

ST1- unadjusted MNLR

## Data Availability

BRFSS 2018 is a publically available database for anyone to access. https://www.cdc.gov/brfss/annual_data/annual_2018.html

