## Supplemental Table 1 for "US women screen at lower rates for both cervical and colorectal cancers than a single cancer"

Supplemental Figure. **CONSORT DIAGRAM**

**Exclusions**

Hysterectomy: N=19,011

Endometrial cancer: N=1

Cervical cancer : N=29

Colon cancer : N=17

Rectal cancer: N=6

**Analysis Group**

**N=40,511**

Women 50-65 years old:

**N=67,473**

**N=48,209**

**Exclusions**

Incomplete data on screening outcomes: N=7,898
