## Supplementary material for "US women screen at lower rates for both cervical and colorectal cancers than a single cancer": ST1- unadjusted MNLR

Supplement Table 1. Unadjusted Multinomial regression

|  | **Referent Outcome: Neither Screen** | | | **Referent Outcome: Dual Screens** | | **Referent Outcome:**  **Cervical Only** |
| --- | --- | --- | --- | --- | --- | --- |
|  | **Dual Screens** | **Cervical Only** | **CRC Only** | **Cervical Only** | **CRC Only** | **CRC Only** |
|  | **OR (95% CI)** | **OR (95% CI)** | **OR (95% CI)** | **OR (95% CI)** | **OR (95% CI)** | **OR (95% CI)** |
| **Age** |  |  |  |  |  |  |
| 50 to 54 | *Ref* | *Ref* | *Ref* | *Ref* | *Ref* | *Ref* |
| 55 to 59 | ***1.29 (1.07, 1.56)*** | ***0.55 (0.45,0.68)*** | ***1.68 (1.24, 2.29)*** | ***0.43 (0.37, 0.49)*** | ***1.30 (1.01, 1.69)*** | ***3.05 (2.31, 4.01)*** |
| 60 to 64 | ***1.48 (1.24, 1.78)*** | ***0.45 (0.37, 0.56)*** | ***2.74 (2.04, 3.70)*** | ***0.31 (0.27, 0.35)*** | ***1.85 (1.44, 2.39)*** | ***6.04 (4.60, 7.93)*** |
| **Race** |  |  |  |  |  |  |
| White | *Ref* | *Ref* | *Ref* | *Ref* | *Ref* | *Ref* |
| Black | ***1.28 (0.98, 1.67)*** | ***1.33 (1.01, 1.76)*** | 1.20 (0.77, 1.86) | 1.04 (0.88, 1.22) | 0.93 (0.64, 1.36) | 0.90 (0.61, 1.33) |
| Other ^a^ | 0.73 (0.49, 1.09) | 0.76 (0.49, 1.17) | ***0.35 (0.21, 0.60)*** | 1.04(0.77, 1.40) | ***0.49 (0.32,0.74)*** | ***0.47 (0.29, 0.74)*** |
| Hispanic | 0.80 (0.64, 1.00) | ***1.63 (1.25, 2.12)*** | 0.96 (0.62, 1.51) | ***2.04 (1.67, 2.50)*** | 1.21 (0.80,1.83) | ***0.59 (0.38, 0.91)*** |
| **Education** |  |  |  |  |  |  |
| Less than High School | *Ref* | *Ref* | *Ref* | *Ref* | *Ref* | *Ref* |
| High School Graduate | ***1.54 (1.20, 1.98)*** | 0.90 (0.67, 1.21) | 1.14 (0.76, 1.72) | ***0.58 (0.46, 0.75)*** | 0.74 (0.51, 1.08) | 1.27 (0.84, 1.91) |
| Attended college/Tech School | ***2.62 (2.03, 3.37)*** | 1.29 (0.96, 1.74) | 1.42 (0.96, 2.11) | ***0.49 (0.39, 0.63)*** | ***0.54 (0.38, 0.78)*** | 1.10 (0.75, 1.62) |
| Graduated college/Tech school | ***4.84 (3.67, 6.38)*** | ***1.96 (1.43, 2.69)*** | ***1.64 (1.09, 2.45)*** | ***0.41 (0.32, 0.52)*** | ***0.34 (0.24, 0.48)*** | 0.83 (0.57, 1.22) |
| **Income** |  |  |  |  |  |  |
| <$50K | *Ref* | *Ref* | *Ref* | *Ref* | *Ref* | *Ref* |
| >$50K | ***3.30 (2.79, 3.90)*** | ***2.07 (1.72, 2.48)*** | ***1.60 (1.24, 2.07)*** | ***0.63 (0.56, 0.70)*** | ***0.49 (0.39, 0.60)*** | ***0.78 (0.62, 0.97)*** |
| **Marital Status** |  |  |  |  |  |  |
| Married or partnered | *Ref* | *Ref* | *Ref* | *Ref* | *Ref* | *Ref* |
| Single ^b^ | ***0.56 (0.48, 0.66)*** | ***0.68 (0.57,0.81)*** | ***0.78 (0.61, 0.99)*** | ***1.21 (1.08, 1.35)*** | ***1.38 (1.13,1.69)*** | 1.14 (0.92, 1.42) |
| **Occupational Status** |  |  |  |  |  |  |
| Employed | *Ref* | *Ref* | *Ref* | *Ref* | *Ref* | *Ref* |
| Unemployed, looking ^c^ | ***0.56 (0.41, 0.78)*** | ***0.64 (0.46, 0.88)*** | ***0.81 (0.51, 1.28)*** | 1.12 (0.87, 1.45) | 1.43 (0.95, 2.17) | 1.27 (0.83, 1.94) |
| Unemployed, not looking ^c^ | 0.99 (0.83, 1.19) | ***0.76 (0.60, 0.96)*** | 1.22 (0.92, 1.63) | ***0.76 (0.64, 0.91)*** | 1.23 (0.97, 1.57) | ***1.61 (1.22, 2.13)*** |
| Unable to Work | ***0.64 (0.52,0.79)*** | ***0.64 (0.50, 0.81)*** | ***1.52 (1.10, 2.10)*** | 0.99 (0.84, 1.16) | ***2.36 (1.81, 3.08)*** | ***2.39 (1.78, 3.20)*** |
| **Location** |  |  |  |  |  |  |
| Urban | *Ref* | *Ref* | *Ref* | *Ref* | *Ref* | *Ref* |
| Rural | ***0.64 (0.52, 0.78)*** | ***0.76 (0.61, 0.95)*** | 0.97 (0.72, 1.30) | ***1.20 (1.04, 1.39)*** | ***1.52 (1.20, 1.94)*** | 1.27 (0.98, 1.65) |
| **Chronic conditions ^d^** |  |  |  |  |  |  |
| Cardiac | ***0.64 (0.48, 0.87)*** | ***0.59 (0.42,0.83)*** | 1.11 (0.74, 1.65) | 0.92 (0.74, 1.14) | ***1.72 (1.26, 2.35)*** | ***1.87 (1.33, 2.64)*** |
| Stroke | 0.96 (0.64, 1.42) | 0.80 (0.52, 1.23) | 0.90 (0.54, 1.51) | 0.84 (0.64, 1.10) | 0.94 (0.63, 1.39) | 1.12 (0.73, 1.72) |
| Lung | 0.95 (0.80, 1.14) | 0.89 (0.70, 1.13) | 1.15 (0.87, 1.53) | 0.93 (0.77, 1.12) | 1.21 (0.95, 1.54) | 1.30 (0.97, 1.74) |
| Arthritis | ***1.51 (1.30, 1.76)*** | 0.99 (0.83, 1.18) | ***2.27 (1.80, 2.86)*** | ***0.65 (0.58, 0.74)*** | ***1.50 (1.24, 1.81)*** | ***2.29 (1.85, 2.82)*** |
| Kidney | 0.84 (0.57, 1.23) | ***0.56 (0.36, 0.87)*** | 0.76 (0.44, 1.30) | ***0.66 (0.49, 0.91)*** | 0.90 (0.59, 1.39) | 1.36 (0.83, 2.23) |
| Diabetes | 1.04 (0.83, 1.29) | ***1.18 (0.89, 1.58)*** | ***1.72 (1.24, 2.38)*** | ***1.66 (1.27, 2.15)*** | ***1.66 (1.27, 2.15)*** | ***1.46 (1.05, 2.01)*** |
| Depression | 1.16 (0.99, 1.37) | 0.97 (0.79, 1.20) | ***1.56 (1.21, 2.01)*** | ***0.84 (0.72, 0.98)*** | ***1.34 (1.08, 1.65)*** | ***1.60 (1.25, 2.05)*** |
| Skin Cancer | ***1.41 (1.01, 1.95)*** | 0.80 (0.56, 1.14) | 1.11 (0.73, 1.67) | ***0.57 (0.48, 0.68)*** | 0.79 (0.60, 1.04) | ***1.38 (1.01, 1.88)*** |
| Other Cancer | ***3.03 (2.26, 4.06)*** | ***2.16 (1.54, 3.03)*** | ***3.14 (2.07, 4.77)*** | ***0.71 (0.58, 0.88)*** | 1.04 (0.75, 1.43) | ***1.45 (1.01, 2.09)*** |

*Significant results are shown in* ***bold italic*** *font.*

^b^ single includes divorced, widowed, separated and never married.

^c^ “Unemployed, not looking” includes Homemaker, Student and Retired”, “Unemployed, looking” includes out of work over a year and out of work less than a year.

^a^ Other means American Indian, Native Hawaiian, Asian, Multiracial and ‘Other’

^d^ Persons may have more than one condition or cancer

***Screening compared to neither screening***

Compared to neither screening, significant predictors of being up-to-date for **dual screening** were older age, being Black compared to White, having more education, having more income, having arthritis, and having a past skin or any kind of cancer. Alternatively, those who were looking for employment, unable to work, living in a rural area, and having past cardiac disease were the least likely to have the dual screens.

Compared to neither screening, significant predictors of being up-to-date for **only the cervical cancer screening** were women of Black or Hispanic races compared to White, having graduated from college, having more income and having a history of diabetes or another cancer. Older women, being single, unemployed for any reason and unable to work, living in a rural area, having past cardiac or kidney disease, were less likely to have only cervical cancer screening.

Compared to neither screening, significant predictors of being up-to-date for **only CRC screening** were older age, having more education, having more income, being unable to work, and having had arthritis, diabetes, depression or other cancers. Those least likely to have only CRC screening were single women and those unemployed but looking for a job.

***No or some screening compared to dual screening***

Compared to having dual screening, significant predictors of only cervical cancer screening were women who were Hispanic compared to White, single, living in rural area or had a past history of diabetes. Women who were older, had higher education, higher income, were unemployed and had a past history of arthritis, kidney disease, depression, skin and other cancers were less likely to screen for **cervical cancer alone**.

Furthermore, compared to having dual screening, significant predictors of **a CRC screen alone** were women who were older, single, unable to work, lived in a rural area, had past cardiac, arthritis, diabetes or depression. Factors that make CRC screening alone, compared to being up-to-date for both screens less likely were being either American Indian, Native Hawaiian, Asian, or multi-racial, having higher education, and higher income.

***CRC screening alone compared to cervical cancer screening alone***

Women who were older, unemployed, unable to work, had a past history of cardiac, arthritis, diabetes, depression, skin cancer or other cancers were significantly more likely to have a **single CRC cancer screen compared to a cervical cancer screen**. On the other hand, women who were Hispanic, American Indian, Native Hawaiian, Asian, or multi-racial compared to White, and had a higher income were more likely to have **a single cervical cancer screen rather than a CRC screen.**
